# Kinetics of immune responses to the AZD1222/Covishield vaccine with varying dose intervals in Sri Lankan individuals

**DOI:** 10.1101/2021.10.27.21265561

**Authors:** Chandima Jeewandara, Inoka Sepali Aberathna, Laksiri Gomes, Pradeep Darshana Pushpakumara, Saubhagya Danasekara, Dinuka Guruge, Thushali Ranasinghe, Banuri Gunasekera, Achala Kamaladasa, Heshan Kuruppu, Gayasha Somathilake, Osanda Dissanayake, Nayanathara Gamalath, Dinithi Ekanayake, Jeewantha Jayamali, Deshni Jayathilaka, Anushika Mudunkotuwa, Michael Harvie, Thashmi Nimasha, Ruwan Wijayamuni, Lisa Schimanski, Pramila Rijal, Tiong K. Tan, Tao Dong, Alain Townsend, Graham S. Ogg, Gathsaurie Neelika Malavige

## Abstract

**Background:** To understand the kinetics of immune responses with different dosing gaps of the AZD1222 vaccine, we compared antibody and T cell responses in two cohorts with two different dosing gaps.

**Methods:** Antibodies to the SARS-CoV-2 virus were assessed in 297 individuals with a dosing gap of 12 weeks, sampled at 12 weeks post second dose (cohort 1) and in 77 individuals with a median dosing gap of 21.4 weeks (cohort 2) sampled 6 weeks post second dose. ACE2 receptor blocking antibodies (ACE2R-Abs), antibodies to the receptor binding domain (RBD) of the virus and variants of concern (VOC) and ex vivo T cell responses were assessed in a sub cohort.

**Results:** All individuals (100%) had SARS-CoV-2 specific total antibodies and 94.2% of cohort 1 and 97.1% of cohort 2 had ACE2R-blocking Abs. There was no difference in antibody titres or positivity rates in different age groups in both cohorts. The ACE2R-blocking Abs (p<0.0001) and antibodies to the RBD of the VOCs were significantly higher in cohort 2, compared to cohort 1. 41.2% to 65.8% of different age groups gave a positive response by the haemagglutination assay to the RBD of the ancestral virus and VOCs in cohort 1, while 53.6% to 90% gave a positive response in cohort 2. 17/57 (29.8%) of cohort 1 and 17/29 (58.6%) of cohort 2 had ex vivo IFNγ ELISpot responses above the positive threshold. The ACE2R-blocking antibodies and ex vivo IFNγ ELISpot responses at 12 weeks post-first dose, significantly correlated with levels 12 weeks post second dose (Spearman’s r=0.46, p=0.008) and (Spearman’s r=0.71, p<0.0001) respectively.

**Conclusions:** Both dosing schedules resulted in high levels of antibody and T cell responses post vaccination, although those with a longer dosing gap had a higher magnitude of responses, possibly as immune responses were measured 6 weeks post second dose compared to 12 weeks post second dose.

## Background

The ChAdOx1 nCoV-19 vaccine (AZD1222) vaccine, is a non-replicating chimpanzee adenovirus vector vaccine, containing the sequence for the spike protein of the ancestral SARS-CoV-2 virus ^1^. Although the vaccine was initially administered with a dosing gap of 4 weeks between the two doses, subsequently the dosing gap was increased to 12 weeks, as it was shown to increase the efficacy of the vaccine ^2^. However, many countries increased the dosing gap to 16 weeks in order to administer a single dose to a larger population ^3 4^ and also due to the delay in obtaining adequate vaccines for administering the second dose on time ^5^. It was later shown that an increase in the gap between the two doses beyond 12 weeks, and even up to 45 weeks, resulted in higher antibody titres after the second dose of the vaccine ^6^.

AZD1222 was the first vaccine to be rolled out in Sri Lanka, with immunization of the health care workers. However, after it was rolled out to the public, due to the delay in obtaining the second dose, most individuals received their second dose 20 weeks after obtaining their first dose. We showed that at ≥ 16 weeks post-immunization with a single dose of AZD1222, 93.7% of those >70 years had SARS-CoV-2 specific antibodies, although ACE2 receptor blocking antibodies (those that correlate with neutralizing antibodies) were significantly less in those >60 years of age ^5^. However, robust memory T cell and B cell responses were seen in over 90% of individuals. Although it has been shown that an increase in the gap between the two doses subsequently led to higher antibody titres ^6^, there are limited data in the differences in dosing gaps on antibody responses to SARS-CoV-2 variants of concern (VOCs), differences in antibody responses in different age groups, those with comorbidities and also the influence of the dosing gap on T cell responses.

Currently many studies have shown that there is waning of immunity with many of the COVID-19 vaccines following the second dose ^7-9^. Due to an increased rate of breakthrough infections, some which led to hospitalization and death, due to waning of immunity, a booster dose is recommended to elderly individuals and the immunocompromised by many authorities ^10 11^. While waning of antibody levels alone does not necessarily imply waning of efficacy^12^ and protection, it is important to find out if different dosing schedules lead to differences in the quality and quantity of immune responses and thus, an impact of the duration of immunity. In order to answer these questions, we compared the immune responses of two cohorts of Sri Lankan individuals who received the AZD1222 vaccine at 12-week dosing intervals and another cohort who received the vaccine at a median of 21.1 weeks dosing interval. We also investigated the kinetics of antibody and T cell responses in the first cohort (12-week dosing interval) ^13^, who consisted of health care workers, in order to find out if there was waning of immunity.

## Methods

336 HCWs (cohort 1) who received their first dose of AZD1222/Covishield vaccine between the 29^th^ January to 5^th^ of February 2021 and their second dose between 23^rd^ of April to 5^th^ of May were included in the study (12 weeks interval between the two doses) 3 months after receiving the second dose of the vaccine (6 months after the first dose). To compare the influence of gap between the two doses on the immunogenicity of the vaccine, another cohort of individuals (n=88) in the community (cohort 2), who received their first dose between the 15^th^ of February to 4^th^ March 2021 and the second dose between 1^st^ of June to 4^th^ August 2021 were included in the study 6 weeks after their second dose (17 to 24 weeks interval between the two doses). Demographic and the presence of comorbidities such as diabetes, hypertension, cardiovascular disease and chronic kidney disease was determined by a self-administered questionnaire at the time of recruitment from all participants.

Ethics approval was obtained from the Ethics Review Committee of University of Sri Jayewardenepura.

### Detection of SARS-CoV-2 specific total antibodies

The presence of SARS-COV-2 specific total antibodies (IgM, IgA and IgG) were detected by using the Wantai SARS-CoV-2 Ab ELISA (Beijing Wantai Biological Pharmacy Enterprise, China), which detects antibodies to the receptor binding domain (RBD) of the spike protein. The assay was carried out according to the manufacturer’s instructions and the antibody index (an indirect measure of the antibody titre) was calculated by dividing the absorbance of each sample by the cutoff value, according to the manufacturer’s instructions.

### Surrogate neutralizing antibody test (sVNT) to detect ACE2 receptor blocking antibodies (ACE2R-blocking Abs)

The surrogate virus neutralization test (sVNT), which has been shown to correlate with the presence of neutralizing antibodies was used to measure ACE2R-Abs as previously described according to the manufacturer’s instructions (Genscript Biotech, USA) ^14^. This measures the percentage of inhibition of binding of the RBD to recombinant ACE2 and an inhibition percentage ≥ 25% in a sample was considered as positive for ACE2R-<u>blocking</u> Abs ^15^.

### Haemagglutination test (HAT) to detect antibodies to the receptor binding domain (RBD)

The HAT was carried out as previously described using the B.1.1.7 (N501Y), B.1.351 (N501Y, E484K, K417N) and B.1.617.2 versions of the IH4-RBD reagents ^16^, which included the relevant amino acid changes introduced by site directed mutagenesis. The assays were carried out and interpreted as previously described and a titre of 1:20 was considered as a positive response ^13 17^. The HAT titration was performed using 7 doubling dilutions of serum from 1:20 to 1:1280, to determine presence of RBD-specific antibodies. The RBD-specific antibody titre for the serum sample was defined by the last well in which the complete absence of “teardrop” formation was observed. A titre of 1:20 was considered as a positive response, as previously determined ^17^.

### Assays to determine antibodies to the N protein

Qualitative detection of antibodies to SARS-CoV-2 nucleocapsid (N) antigen in human serum was performed using Elecsys® Anti-SARS-CoV-2 electrochemiluminescence immunoassay (Cat: 09 203 095 190, Roche Diagnostics, Germany) in Cobas e 411 analyzer (Roche Diagnostics, Germany). A Cutoff index (COI) ≥1.0 was interpreted as reactive and COI <1.00 was considered non-reactive as per the kit guidelines.

### Ex vivo and cultures IFNγ ELISpot assays

Ex vivo IFNγ ELISpot assays were carried out using freshly isolated peripheral blood mononuclear cells (PBMC) obtained from 57 individuals. Individuals for T cell assays were recruited from those whom these assays had been carried out at 1 month following the first dose of the vaccine ^18^. Two pools of overlapping peptides named S1 (peptide 1 to 130) and S2 (peptide 131 to 253) covering the whole spike protein (253 overlapping peptides) were added at a final concentration of 10 µM and incubated overnight as previously described ^19 20^. All peptide sequences were derived from the wild-type consensus and were tested in duplicate. 100,000 cells/well were added per well. PHA was included as a positive control and media alone was used as a negative control. Briefly, ELISpot plates (Millipore Corp., Bedford, USA) were coated with anti-human IFNγ antibody overnight (Mabtech, Sweden). The plates were incubated overnight at 37°C and 5% CO_2_. The cells were removed, and the plates developed with a second biotinylated antibody to human IFNγ and washed a further six times. The plates were developed with streptavidin-alkaline phosphatase (Mabtech AB) and colorimetric substrate, the spots were enumerated using an automated ELISpot reader (AID Germany). Background (PBMCs plus media alone) was subtracted and data expressed as number of spot-forming units (SFU) per 10^6^ PBMCs. A positive response was defined as mean±2 SD of the background responses.

### Statistical analysis

The 95% confidence intervals for seropositivity for each age category were calculated using the R software (version 4.0.3) and R-studio (version 1.4.1106). Pearson Chi Square Association tests were performed at a confidence level of 95% using the R software to identify the statistically significant associations of the age categories and the sex of the individuals with seropositivity. Kruskal-Wallis test was used to determine the differences between the levels of antibodies between different age groups. Spearman’s correlation coefficient was used to determine the correlation between antibody, T cell responses and the age of an individual.

## Results

### SARS-CoV-2 total antibody responses in individuals with different dosing intervals

At 3 months since receiving the 2^nd^ dose (6 months after the first dose), all of the 336 (100%) of the HCWs (cohort 1) had SARS-CoV-2 specific total antibodies (IgM, IgG and IgA). Antibodies to the N protein were tested in both cohorts (cohort 1 and cohort 2) to determine if they had been infected during the 6 months since obtaining the first dose and 39/336 (11.6%) were excluded from further analysis due to presence of antibodies to the N protein. Therefore, further analysis was carried out in those who were not found to have natural infection during this period (n=297). There was no significant difference (p-value=0.79) between the antibody titres in the three different age groups (20 to 39, 40 to 59 and >60 years) in this cohort 1 (Figure 1A).

**Figure 1:**
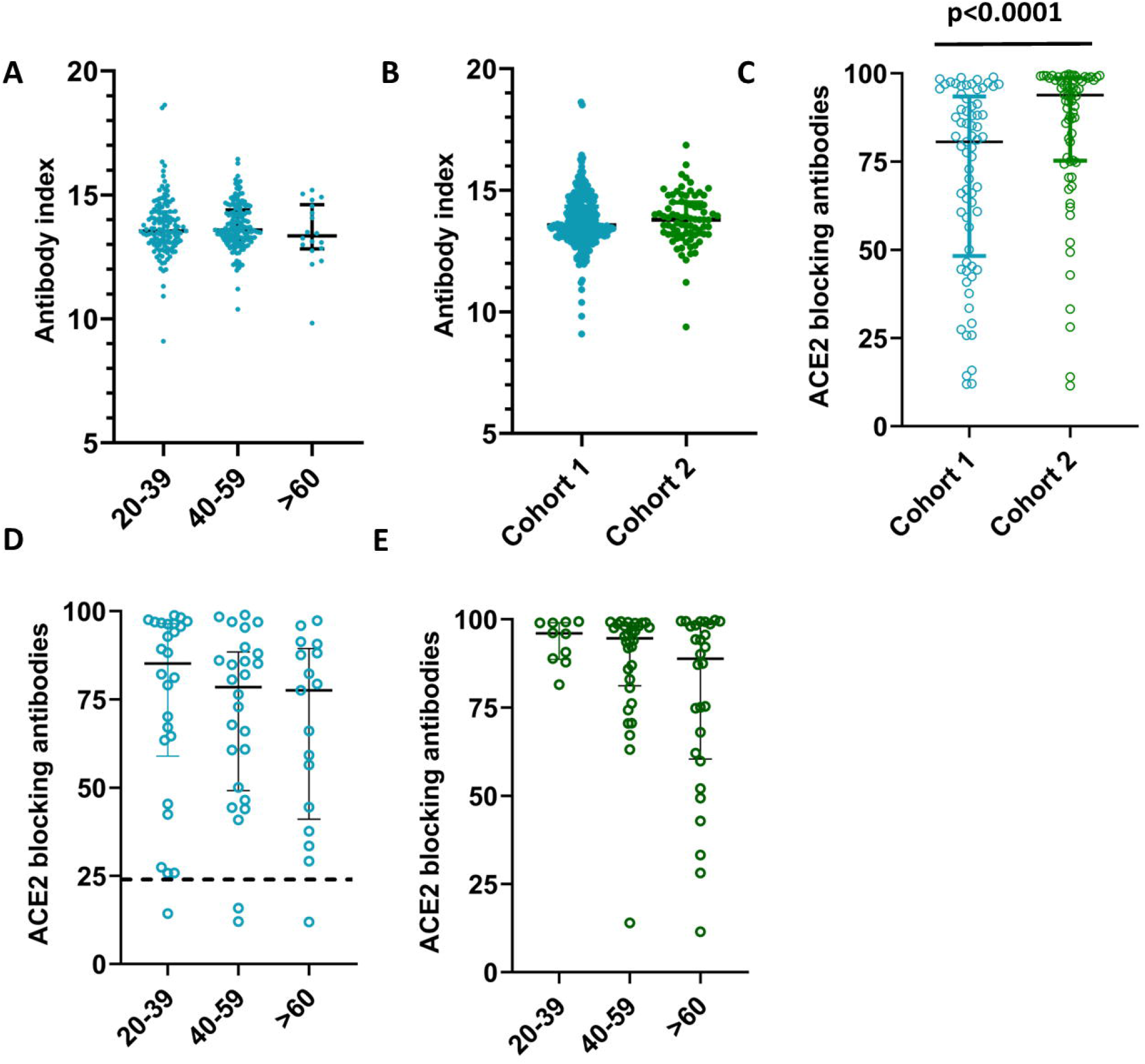
SARS-CoV-2 specific antibodies in individuals of different age groups in cohort 1 and cohort 1. SARS-CoV-2 specific total antibodies were measured in 20-to-39-year old’s (n=129), 40-to-59-year old’s (n=152) and those >60 years of age (n=16) by ELISA in cohort 1, and no significant difference was seen between the age groups (p=0.06) based on the Kruskal-Wallis test (A). SARS-CoV-2 specific total antibodies were measured in cohort 1 (n=297) and cohort 2 (n=77) and no significant different was seen based on the Mann-Whitney test (B). The ACE2 receptor blocking antibodies were measured by the surrogate virus neutralizing test in cohort 1 (n=69) and cohort 2 (n=70), which was significantly different (p<0.0001) (C). The ACE2 receptor blocking antibodies were measured in cohort 1 in 20-to-39-year old’s (n=26), 40-to-59-year old’s (n=26) and >60-year old’s (n=17) and no significant difference was seen (p=0.41) between the age groups based on the Kruskal-Wallis test (D). The ACE2 receptor blocking antibodies were also in cohort 2 in 20-to-39-year old’s (n=10), 40-to-59-year old’s (n=32) and >60-year old’s (n=28) and no significant difference was seen (p=0.30) between the age groups based on the Kruskal-Wallis test (E). All tests were two-tailed. The lines indicate the median and the inter quartile range. All data points of cohort 1 is shown in blue and in cohort 2 in green.

The second cohort was recruited at 6 weeks since receiving the 2^nd^ dose (6 to 7 months after the first dose) and the mean gap between the two doses in this cohort was 21.14 weeks (SD ±1.95 weeks). Of this cohort, 11/88 (12.5%) had antibodies to the N protein and were considered as having been infected with the SARS-CoV-2 virus. All of the 77 individuals (100%) of this cohort (cohort 2) also had SARS-CoV-2 specific total antibodies. There was no significant difference between the antibody titres in cohort 1 compared to cohort 2 (p-value=0.3488) (Figure 1B). As seen with cohort 1, there was no significant difference (p-value=0.5716) between the antibody titres in the three different age groups (20 to 39, 40 to 59 and >60 years) in cohort 2. In both cohorts, there was no significant difference (p-values=0.96, 0.99) in the seropositivity rates or the antibody levels (indicated by antibody indices) in those with comorbidities, compared to those who did not have comorbidities.

### SARS-CoV-2 specific ACE2-receptor blocking antibodies (ACE2R-<u>blocking</u> Abs) in the two cohorts with different dosing intervals

The surrogate neutralizing antibody test (sVNT) that measures ACE2R-<u>blocking</u> Abs was carried out in a subset of individuals of cohort 1 (n=69) and cohort 2 (n=70). In cohort 1 (those with a 12-week gap between the two doses), 65/69 (94.2 %) gave a positive result for the presence of ACE2R-<u>blocking</u> Abs, while 68/70 (97.1%) in cohort 2 gave a positive response. The positivity rates were found to be significantly different higher in cohort 1 than cohort 2 (Mann-Whitney U = 1472, p<0.0001) and cohort 2 had significantly higher titres (median 93.7, IQR 75.3 to 98.7 % of inhibition) compared to cohort 1 (median 80.6, IQR 48.3 to 93.5 % of inhibition (Figure 1C). The ACE2R-<u>blocking</u> Ab levels in the three different age groups in cohort 1 and cohort 2 are shown in table 1. There was no significant different between the ACE2R-<u>blocking</u> Abs in different age groups in cohort 1 (p=0.41) (Figure 1D), or cohort 2 (p=0.30) (Figure 1E).

**Table 1:** ACE2R-Ab positivity rates and the levels in different age groups in cohort 1 and cohort 2.

| Age groups | Presence of ACE2R-<u>blocking</u><br>Abs | % of inhibition as given by the<br>sVNT assay (median, IQR) |
| --- | --- | --- |
| Cohort 1 |  |  |
| 20 to 39 (n=26) | 25 (96.1%) | (85.2, 96.4-63.7=32.7) |
| 40 to 59 (n=26) | 24 (92.3 %) | (78.5, 87.5-52.7=34.8) |
| >60 (n=17) | 16 (94.1 %) | (77.6, 88.1-44.4=43.7) |
| Cohort 2 |  |  |
| 20 to 39 (n=10) | 10 (100%) | (96.0, 99.0-89.3=9.6) |
| 40 to 59 (n=32) | 31 (96.9%) | (94.6, 98.1-82.4=15.7) |
| >60 (n=28) | 27 (96.4%) | (88.8, 98.4-61.5=36.9) |

### Hemagglutination test (HAT) to detect antibodies to the receptor binding domain of SARS-CoV-2 and its variants of concern (VOCs)

The HAT assay was carried out to measure positivity rates and the antibody titres to the ancestral strain (WT), and the VOCs B.1.1.7, B.1.351 and B.1.617.2 in cohort 1 (n=69) and in cohort 2 (n=70). These are the same individuals in whom ACE2 receptor blocking antibodies were measured. The proportion of individuals who gave a positive result for the WT and the VOCs is shown in table 2. As determined by the Friedman test, in cohort 1 the HAT titres for different VOCs were not found to be significantly different in any age group; 20 to 39 years (p=0.068), 40 to 59 years (p=0.658) and the >60 year age group (p=0.251) (Figure 2A). The lowest titres were seen for B.1.351, while those who were in the 40 to 59 age group had low titres for B.1.617.2. In cohort 2, again there was no difference in the HAT titres between the WT and the VOCs (Figure 2B).

**Table 2:**
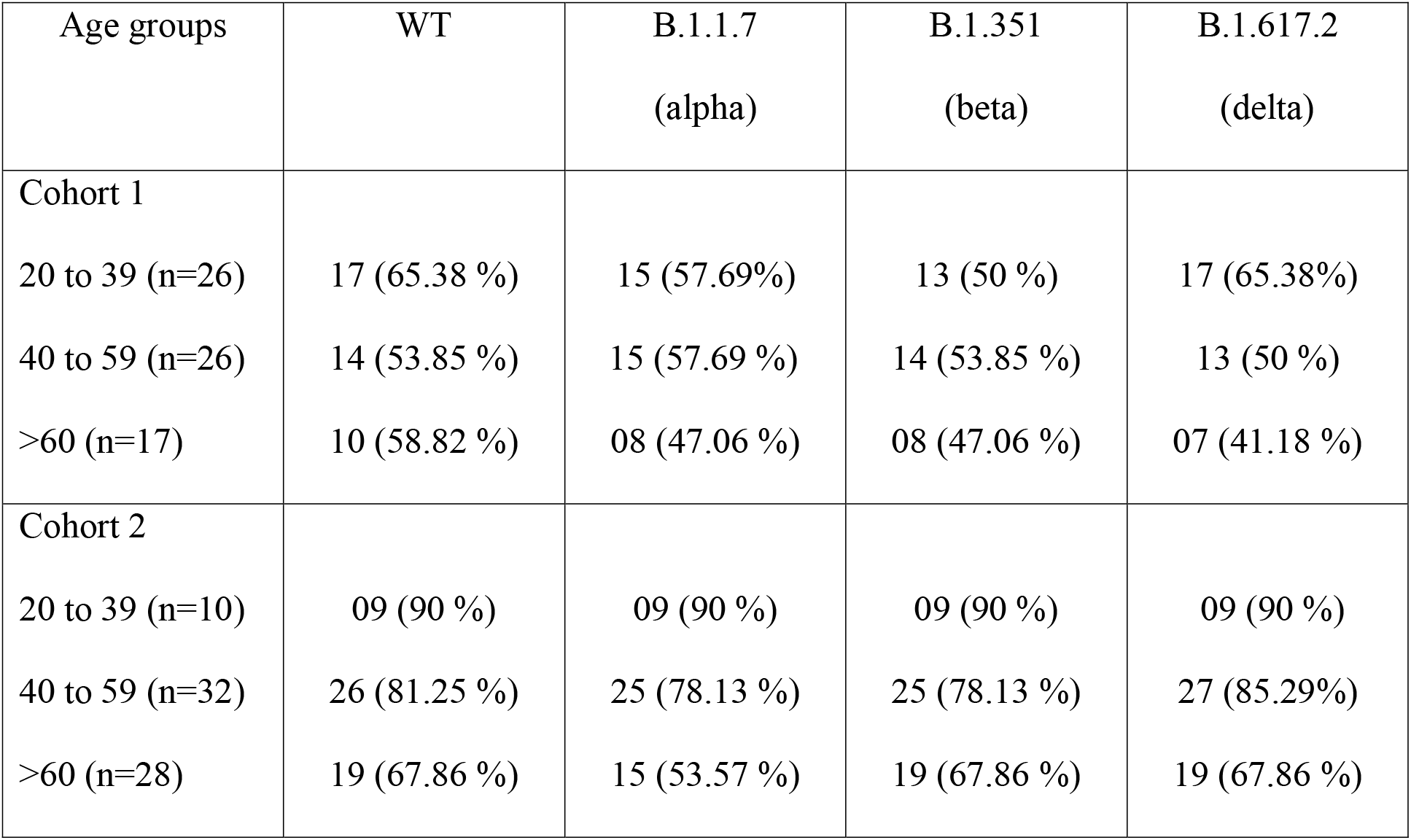
Positivity rates and titres for the WT and SARS-CoV-2 variants of concern, in different age groups in cohort 1 and cohort 2 measured by the haemagglutination assay.

| Age groups | WT | B.1.1.7<br>(alpha) | B.1.351<br>(beta) | B.1.617.2<br>(delta) |
| --- | --- | --- | --- | --- |
| Cohort 1 |  |  |  |  |
| 20 to 39 (n=26) | 17 (65.38 %) | 15 (57.69%) | 13 (50 %) | 17 (65.38%) |
| 40 to 59 (n=26) | 14 (53.85 %) | 15 (57.69 %) | 14 (53.85 %) | 13 (50 %) |
| >60 (n=17) | 10 (58.82 %) | 08 (47.06 %) | 08 (47.06 %) | 07 (41.18 %) |
| Cohort 2 |  |  |  |  |
| 20 to 39 (n=10) | 09 (90 %) | 09 (90 %) | 09 (90 %) | 09 (90 %) |
| 40 to 59 (n=32) | 26 (81.25 %) | 25 (78.13 %) | 25 (78.13 %) | 27 (85.29%) |
| >60 (n=28) | 19 (67.86 %) | 15 (53.57 %) | 19 (67.86 %) | 19 (67.86 %) |

**Figure 2:**
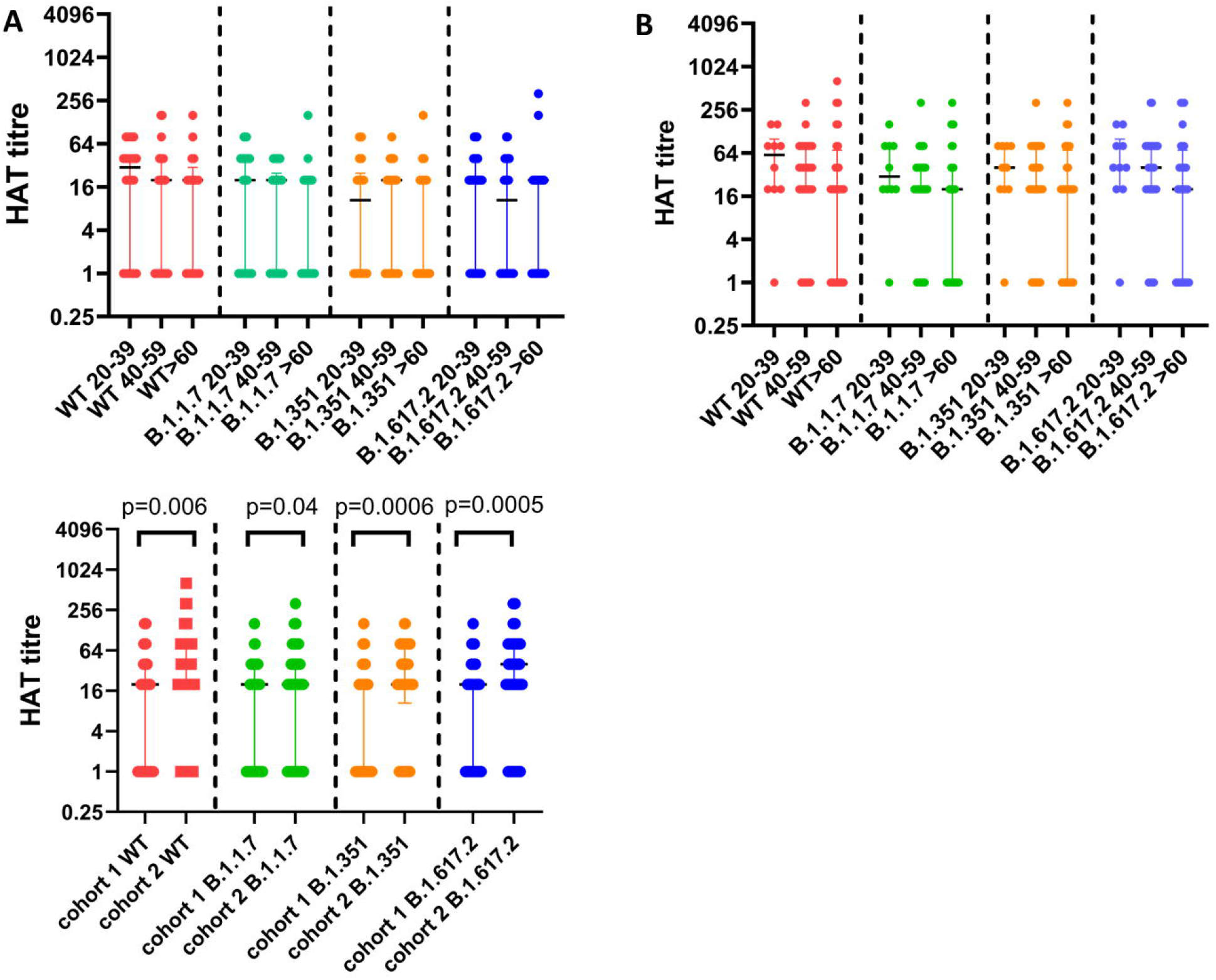
Antibodies to the receptor binding domain (RBD) of the ancestral SARS-CoV-2 virus (WT) and variants of concern in cohort 1 and 2. Antibodies to the RBD of the WT and VOCs were measured in cohort 1 in 20-to-39-year old’s (n=26), 40-to-59-year old’s (n=26) and >60-year old’s (n=17) and no significant difference was seen between the different age groups for responses to the RBDs of different VOCs (A). Antibodies to the RBD of the WT and VOCs were also measured in cohort 2 in 20-to-39-year old’s (n=10), 40-to-59-year old’s (n=32) and >60-year old’s (n=28) and no significant difference was seen between the different age groups for responses to the RBDs of different VOCs (B). The HAT titres for the WT, B.1.1.7, B.1.351 and B.1.617.2 was compared between the two cohorts. Individuals in cohort 2 had significantly higher HAT titres to the WT and all VOCs analysed by the Mann-Whitney test. All tests were two-tailed. The lines indicate the median and the inter quartile range.

The HAT titres for the WT (p=0.006), B.1.1.7 (p=0.04), B.1.351 (p=0.0006) and B.1.617.2 (p=0.0005) were significantly higher in those in cohort 1 compared to cohort 2 (Figure 2C).

### Ex vivo ELISpot responses in the two cohorts with different dosing schedules

To investigate the T cell responses in these two cohorts, we carried out ex vivo IFNγ ELISpot responses in cohort 1 (n=57) and in cohort 2 (n=29). In cohort 1, the positive threshold was set at 110 SFU/1 million PBMCs and accordingly 17/57 (29.8%) gave a positive response. In cohort 2, the threshold for a positive response was set at 272 SFU/1 million cells. Accordingly, 17/29 (58.6%) gave a positive response. The frequency of ex vivo IFNγ ELISpot responses were significantly higher for the S1 pool of over lapping peptides (p=0.009) and the S2 pool of overlapping peptides (p=0.0006) in cohort 2 compared to cohort 1 (Figure 3).

**Figure 3:**
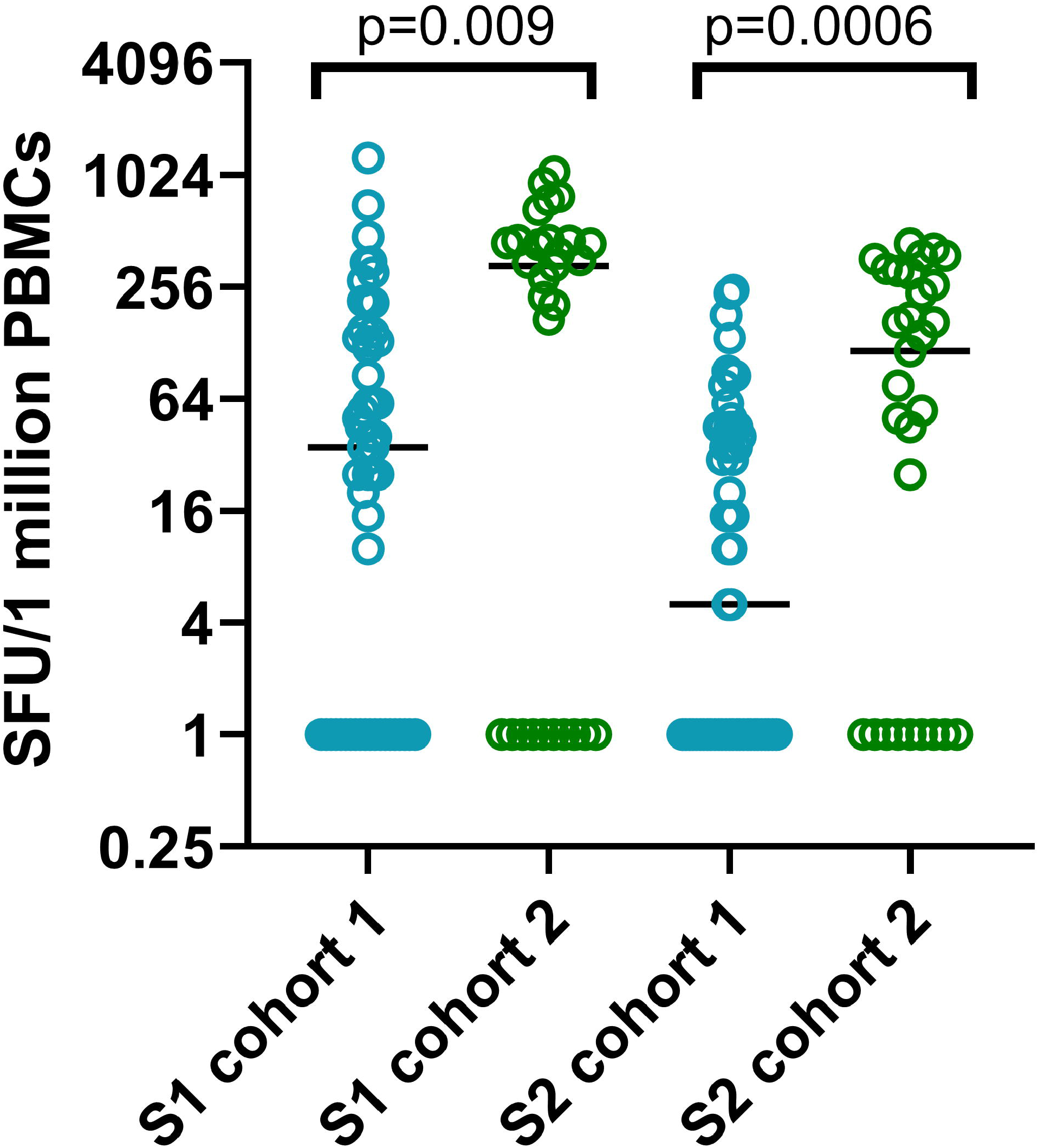
Ex vivo IFNγ ELISpots responses to the overlapping peptides of the spike protein in the two cohorts. Ex vivo IFNγ ELISpots responses were measured to the S1 and S2 overlapping pool of peptides in 57 individuals in cohort 1 (blue) and 29 individuals in cohort 2 (green). The frequency of ex vivo IFNγ ELISpot responses were significantly higher for the S1 pool of over lapping peptides (p=0.009) and the S2 pool of overlapping peptides (p=0.0006) in cohort 2 compared to cohort 1, based on the Mann-Whitney test. All tests were two-tailed.

The responses to the S1 pool of overlapping peptides (median 35, IQR 0 to 132.5 SFU/1 million PBMCs) were significantly higher (p<0.0001) than for the S2 pool of overlapping peptides (median 5, IQR 0 to 45 SFU/1 million PBMCs) in cohort 1. In cohort 2, although the responses to S1 pool was higher (median 330, IQR 0 to 452.5 SFU/1 million PBMCs) than those for S2 (median 115, IQR 0 to 305 SFU/1 million PBMCs), this was not significant (p=0.06).

### Kinetics of antibody responses and T cell responses in cohort 1 over time

The kinetics of antibody and T cell responses could only be studies in cohort 1, as we had data at baseline, 4 weeks after the first dose, 12 weeks after the first dose and 12 weeks after the second dose (6 months after the first dose) for cohort 1. Only 171 individuals (N antibody negative) were included in the analysis for kinetics of the SARS-CoV-2 total antibodies as all four time points were available only in these individuals. Of the 171 individuals, 73 were in the 29 to 39 age group, 87 in the 40 to 59 age group and 11 in the >60 age group. The SARS-CoV-2 total antibodies were significantly higher from the time from obtaining the second dose to 12 weeks later (p<0.0001) (Figure 4A). However, while the SARS-CoV-2 specific total antibodies rose with time in all age groups, this rise was only significant in the 40 to 59 age group (p=0.001), while there was no significant difference in those in the 20 to 39 age group and >60 age group, in the levels from 12 weeks post-first vaccine and 12 weeks post-second vaccine (Figure 4A). In the 20 to 39 age group and the 40 to 59 age group the antibody levels were significantly higher at 12 weeks post second dose (24 months post first dose) than at 4 weeks post first dose (p<0.0001). In the >60 age group, although the levels at 12 weeks post-second dose was significantly higher than that of 4 weeks post first dose (p=0.049), the levels were still lower.

**Figure 4:**
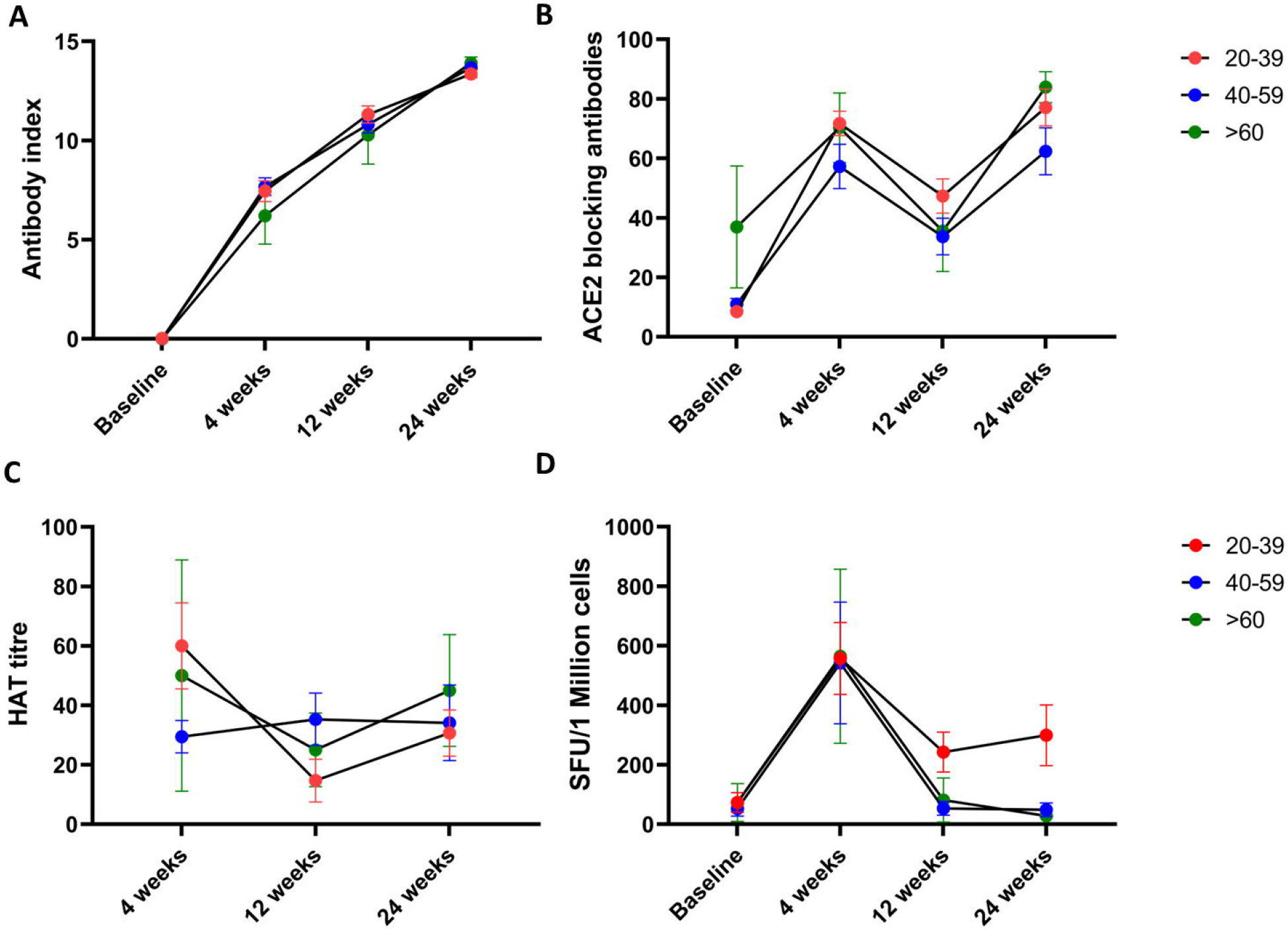
Kinetics of antibody and T cell responses over time in cohort 1 (dosing gap of 12 weeks). SARS-CoV-2 total antibodies were measured in 171 individuals, (73 in the 29 to 39 age group, 87 in the 40 to 59 age group and 11 in the >60 age group), at baseline, 4 weeks after the first dose, 12 weeks post-first dose and 12 weeks post second dose by ELISA (A). ACE2 receptor antibodies were measured by the surrogate virus neutralizing test in 33 individuals (16 in 29 to 30 age group, 12 in 40 to 59 and 5>60 age group (B). Antibodies to the RBD of the WT was measured by the haemagglutination assay test in 40 individuals (C). Ex vivo IFNγ ELISpot responses to the S protein overlapping pool of peptides were measured in 36 individuals, with16 in the 20 to 39 age group, 17 in the 40 to 59 age group and 3 >60 years. There was no difference in responses between 6 to 16 weeks. (D). The lines indicate the mean and the error bars the standard error of the mean. All tests were two-tailed.

The ACE2R-<u>blocking</u> Ab levels were only available in 33 individuals (16 in 29 to 30 age group, 12 in 40 to 59 and 5>60 age group), in cohort 1 for all 4 time points. Using the Friedman test we found that the ACE2R-Ab levels were significantly higher (p<0.0001) at 12 weeks post second dose compared to 12 weeks post first dose (Figure 4B). This rise in ACE2R-<u>blocking</u> Abs were only significantly higher in the 20 to 39 age group (p=0.002), but not in the other two age groups. The antibodies to the RBD of the WT was also assessed over time in this cohort (n=40), and there was no significant difference (p=0.59) in the antibody titres to the RBD of the WT, assessed by the haemagglutination assay from 12 weeks post second dose compared to 12 weeks post first dose (Figure 4C).

The kinetics of ex vivo IFNγ ELISpot responses were assessed in 36 individuals over time (16 in the 20-39 age group, 17 in the 40-59 age group and 3 >60 years). While the ex vivo spike protein specific (overlapping peptide) responses increased with time, there was no significant difference (p>0.99) in responses 12 weeks post second dose compared to 12 weeks post first dose (Figure 4D), in any of the three age groups. Statistical analysis was not carried out between individual age groups as there were only 3 individuals >60 years of age. However, the T cell responses were higher in those in the 20 to 39 age group.

### Association of immune responses to the first dose with those of immune responses following post-second dose of the vaccine

In cohort 1, The antibody levels at 12 weeks post single dose significantly correlated with the SARS-CoV-2 total antibodies at 12 weeks post second dose (Spearmans’r= 0.22, p=0.001). However, in further analysis this correlation was only seen in the 20 to 39 age group (Spearman’d r=0.37, p=0.0003), whereas no significant correlation was seen in other age groups. However, no such correlation was seen in the total antibodies at ≥16 weeks post first dose and 6 weeks post second dose in cohort 2 (Spearmans r=0.03, p=0.82).

In cohort 1 (Spearman’s r=0.46, p=0.008) the ACE2R-blocking antibodies at 12 weeks post-first dose, significantly correlated with levels 12 weeks post second dose. A significant correlation was seen in cohort 2 (Spearman’s r= 0.41, p=0.001) as well in antibody levels at ≥16 weeks post first dose and 6 weeks post second dose. The ex vivo IFNγ ELISpot responses showed the strongest correlation in cohort 1 in 12 weeks post first dose and 12 weeks post second dose (Spearman’s r=0.71, p<0.0001). Data was not available to carry out this analysis for cohort 2.

## Discussion

In this study we have compared the antibody and T cell responses of individuals with two dosing schedules, 6 months after receiving the first dose of the AZD1222 vaccine. In the first cohort with a dosing gap of 12 weeks, individuals were recruited 12 weeks post second dose, while in cohort 2, with a dosing gap of a median 21.4 weeks, individuals were recruited 6 weeks post second dose. We found that all individuals in both cohorts were seropositive and there was no difference in the SARS-CoV-2 total antibodies between the two cohorts. However, cohort 2 had significantly higher levels of ACE2R-<u>blocking</u> Abs and antibodies to the RBD of the WT and VOCs when compared to cohort 1. The high ACE2R-Abs and antibodies to WT in the second cohort could be because they were 6 weeks post second dose compared to cohort 1, which was 12 weeks post second dose. Therefore, it is possible that those in cohort 2 had not entered the contraction phase of the memory response by 6 weeks post-second dose and therefore, had higher T cell and antibody responses. However, both cohorts were 6 months post first dose, and at that time point, those with a longer dosing gap had higher antibody titres. Although we did not assess neutralizing antibodies in this study, ACE2R-<u>blocking</u> Abs and antibodies to the RBD detected by the HAT assay has been shown to significantly correlate with levels of neutralizing antibodies ^14 21^. Since neutralizing antibodies have shown to associate with protection against SARS-CoV-2 virus infection ^22^, from 6 months post first dose, those with a longer dosing interval appear to have a higher level of protection.

We previously showed that the levels of ACE2R-<u>blocking</u> Abs declined from 4 weeks to 12 weeks after a single dose and thereby possibly increasing the susceptibility to infection by 16 weeks^5^. However, 73.9% still had detectable ACE2R-<u>blocking</u> Abs, while robust T and B cell memory responses were seen in >90% at 16 weeks ^5^. Although there are no data regarding the efficacy of a single dose of the vaccine in preventing infection at ≥16 weeks, we found that 12.5% of this cohort with a longer dosing gap had been infected with the virus, with mildly symptomatic or asymptomatic infection. This is not surprising as Sri Lanka had a high number of cases from April to June due to the alpha variant ^23^ and even a higher number of cases with the delta variant from June to end of September due to the delta variant ^24 25^. Therefore, although a large proportion (11/88) had evidence of infection, the vaccine appeared to have induced an adequate immune response to prevent hospitalization. The waning of neutralizing antibody responses has been observed in especially older individuals with time, and therefore, many countries have recommended a booster dose 6 months after the second dose of the vaccine ^10 26^. However, those who had a prolonged dosing interval appear to have higher antibody and T cell responses and therefore, are likely to have higher neutralizing antibody responses for a longer duration from the time they received the first dose. Therefore, the prolonged gap between the two doses may be beneficial in vaccine roll-out by enabling more individuals to receive one dose of a vaccine, but not compromising immunity and possibly delaying giving out booster doses.

Interestingly, we found that while only a significant correlation was seen for the total SARS-CoV-2 specific antibodies between the 12 weeks post first dose and 12 weeks post second dose in 20 to 39 years olds and not in cohort 2, the ACE2R-blocking antibodies post second dose significantly correlated with the values post first dose. A significant correlation for ex vivo IFNγ ELISpot responses was only observed in cohort 1, where a strong correlation was seen between T cell responses at 12 weeks following the first dose when compared to T cell responses 12 weeks following the second dose. Therefore, individuals who have the highest frequency of responses to the first dose appear to also have a higher magnitude of responses to the second dose. Therefore, the level of immune responses before second dose appears to be an important factor determines the magnitude of the responses after the second dose.

Our data show that 12 weeks after post second dose, while 94.2% of those with a 12-week gap had ACE2R-<u>blocking</u> Abs with median antibody titers of 77.6 (% of inhibition). In contrast, we showed that 12 weeks after the second dose of Sinopharm/BBIBP-CorV, only 60.9% of individuals had ACE2R-<u>blocking</u> Abs, with median titres of 35.6% ^9^. Therefore, there appears to be significant differences in the kinetics of the immune responses with time, for different types of vaccines. It would be important to take into account these differences, when decisions regarding when and to whom to administer booster doses are taken, in order to prevent large outbreaks of breakthrough infection. However, although neutralizing antibodies have shown to associate with protection ^22^, and T cells have shown to associate with reduced disease severity ^27^, the correlates of a protective immune response are yet unknown. Therefore, the efficacy of different dosing schedules and for different vaccines in providing durable immunity can only be evaluated by clinical trials.

In summary, we have investigated the immune responses following two dosing schedules of the AZD1222 in Sri Lankan individuals. We found that those who had a wider dosing gap had higher antibody and T cell responses, 6 months post-first dose of the vaccine, when compared to those with a 12-week dosing gap. It would be important to assess the significance of these differences in immune responses based on the dosing gap on vaccine efficacy.

## Data Availability

All data is available in the manuscript and the figures

## Acknowledgement

We are grateful to the Allergy, Immunology and Cell Biology Unit, University of Sri Jayewardenepura; the NIH, USA (grant number 5U01AI151788-02), UK Medical Research Council and the Foreign and Commonwealth Office for support. T.K.T. is funded by the Townsend-Jeantet Charitable Trust (charity number 1011770) and the EPA Cephalosporin Early Career Researcher Fund. A.T. are funded by the Chinese Academy of Medical Sciences (CAMS) Innovation Fund for Medical Science (CIFMS), China (grant no. 2018-I2M-2-002).

